# Impact of banning smoking in cars with children on exposure to second-hand smoke: a natural experiment in England and Scotland

**DOI:** 10.1101/19006353

**Authors:** Anthony A Laverty, Thomas Hone, Philip E. Anyanwu, David Taylor Robinson, Frank de Vocht, Christopher Millett, Nicholas S Hopkinson

## Abstract

A ban on smoking in cars with children was implemented in April 2015 in England and December 2016 in Scotland. With survey data from both countries (N_England_=3,483-6,920 and N_Scotland_=232-319), we used this natural experiment to assess the impact of the ban using a difference-in-differences approach. We conducted logistic regression analyses within a Difference-in-Difference framework and adjusted for age, sex, a marker of deprivation and survey weighting for non-response. Among children aged 13-15 years, self-reported levels of regular exposure to smoke in cars were 3.4% in 2012, 2.2% in 2014 and 1.3% in 2016 for Scotland and 6.3%, 5.9% and 1.6% in England. The ban was associated with a 73% reduction (95%CI -59%, -81%) in self-reported exposure to tobacco smoke among children.

## INTRODUCTION

Exposure to second-hand tobacco smoke is a significant cause of illness in children and particularly affects more disadvantaged groups [1]. Exposure of children to smoking inside cars is especially concerning due to the very high concentrations that accumulate in these enclosed spaces and its association with a greater risk of child smoking uptake [2][3]. Both smoking uptake and levels of child exposure to smoke in cars are socially patterned, and as such serve as mechanisms which sustain health inequality [4][5][6].

One policy response has been to ban smoking in private vehicles with children present. Evaluations of the impact of banning smoking in cars with children are scarce and present a mixed picture [7][8]. A ban on smoking in private vehicles with anyone ≤18 years present came into effect on 1^st^ October 2015 in England, and on 5^th^ December 2016 in Scotland. This difference in timing provides a natural experiment to evaluate the impact of the policy.

## METHODS

Data for England came from the Smoking, Drinking and Drug Use surveys and for Scotland from the Scottish Health Surveys in 2012, 2014 and 2016 (further details in Online Supplementary Material). We restricted the sample to children aged 13-15, as children aged 11 and 12 years old had exposure reported by caregivers in Scotland, which is likely to lead to under-recording.

Our primary exposure was child-reported regular exposure to smoking inside cars (**Table 1**). For England, we categorised responses of *every day or most days* or *once or twice a week* as “regular exposure.” In Scotland children were asked “Are you regularly exposed to other people’s tobacco smoke in any of these places?” (Responses: yes/no for a range of locations including “cars/vehicles”). We also included data on age, sex and a marker of deprivation, harmonised between years and countries as set out in **Table 1**.

**Table 1:** Key characteristics of data sources.

| Country | England | Scotland |
| --- | --- | --- |
| Data name | Smoking Drinking Drug Use Survey (SDDU) | Scottish Health Survey |
| Years and interview dates | **2012:** September 2012 – December 2012<br>**2014:** September 2014 – December 2014<br>**2016:** September 2016 – January 2017 | **2012:** January 2012 – December 2012<br>**2014:** January 2014 – February 2015<br>**2016:** January 2016 – January 2017 |
| Individuals included | **2012:** 4,915<br>**2014:** 3,483<br>**2016:** 6,920 | **2012:** 319<br>**2014:** 271<br>**2016:** 232 |
| Age range | 11 - 15 years | 13 - 17 years |
| Exposure question | **2012:** Two separate questions: *“In the past year, how often were you in your family’s car [OR] someone else’s car with somebody smoking?”* (responses: *Every day or most days; Once or twice a week; Once or twice a month; Less often than once a month; Never in the past year; Don’t know*).<br>**2014 & 2016:** In the past year, how often were you in a car with somebody smoking? This could be your family’s car or someone else’s car. (responses: *Every day or most days; Once or twice a week; Once or twice a month; Less often than once a month; Never in the past year; Don’t know*).<br>In this study ‘every day or most days’ classed as “regular exposure” | **2012 – 2016:** “Are you regularly exposed to other people’s tobacco smoke in any of these places?” (responses: yes/no for in cars/vehicles etc) |
| Deprivation marker | **2012 & 2014:** child reported receipt of free school meals (FSM)<br>**2016:** Family Affluence Scale (low, middle, high). Harmonised so low group equivalent to receiving FSM | **2012 – 2016:** Scottish index of Multiple deprivation in five groups.<br>Harmonised so most deprived group comparable to receiving FSM or being in lowest affluence band |

We used survey-weighted logistic regression to assess changes in exposure over time using differences-in-differences analysis which is commonly used for policy evaluation since it controls for all time-invariant differences between the intervention and comparison populations [9]. We modelled a linear trend for time individually for both Scotland and England individually and a binary variable for 2016 in England as the one post-intervention data point, interacted with time.

We conducted using data from England only (ages 11-15) which make use of more granular exposure data to conduct analyses of ever, monthly, and regular exposure.

## RESULTS

There were 15,318 responses in England and 822 in Scotland (**Appendix Table 1)**. Self-reported regular exposure to smoke in cars were 3.4% in 2012, 2.2% in 2014 and 1.3% in 2016 for Scotland and 6.3%, 5.9%, and 1.6% in England (**Figure 1)**.

**Figure 1:**
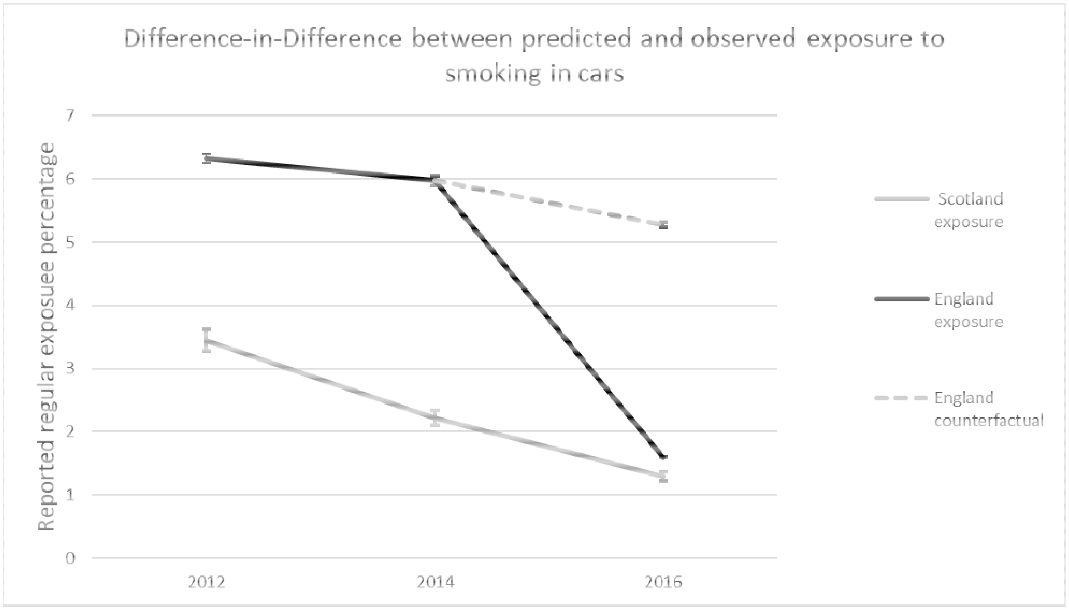
Percentages of children reported regular exposure in England and Scotland with and without policy implementation.

Implementation of the smoke-free policy in England was associated with a 73% reduction in the percentage of children self-reporting exposure to smoke in cars compared to trends in Scotland (AOR=0.27, 95%CI 0.19-0.41) (**Table 2)**. Children in England were more likely to report exposure than those in Scotland (AOR=2.35, 95%CI 1.28-4.30). Girls were more likely to report exposure than boys (AOR=1.61, 95%CI 1.34-1.93), as were those in the deprived group compared with the rest of the sample (AOR=1.98, 95%CI 1.61-2.43).

**Table 2:** Results from logistic regression Difference in Difference analyses of impact of policy implementation on self-reported exposure to smoking in vehicles.

|  | AOR | Lower CI | Upper CI |
| --- | --- | --- | --- |
| Policy Implementation | 0.27 | 0.19 | 0.41 |
| Scotland trend | 0.77 | 0.42 | 1.42 |
| England Trend | 1.01 | 0.82 | 1.21 |
| Scotland baseline | ref | ref | ref |
| England baseline | 2.35 | 1.28 | 4.30 |
| Age 13 years | ref | ref | ref |
| Age 14 years | 1.01 | 0.80 | 1.28 |
| Age 15 years | 1.22 | 0.99 | 1.51 |
| Boys | ref | ref | ref |
| Girls | 1.61 | 1.34 | 1.93 |
| Not deprived group | ref | ref | ref |
| Deprived group | 1.98 | 1.61 | 2.43 |

AOR = Adjusted Odds Ratio, CI = Confidence Interval

Analyses within England only, using the wider age range of 11 to 15 years, identified lower levels of reported exposure after policy implementation than predicted by preceding trends, ranging from AOR 0.75 (95%CI 0.62-0.90 for ever exposure to AOR 0.26 (95%CI 0.18-0.36)) for regular exposure **(Appendix Table 2)**.

## DISCUSSION

The main outcome of the study is that child-reported exposure to tobacco smoke in cars fell following the 2015 introduction of the ban in England, a finding made more robust by the comparison with Scotland where the policy was introduced the following year. We also show that this exposure remains more common in children from more deprived groups, which serves as a reminder of the socially patterned risks of smoking. Our findings provide support for introducing this policy in other jurisdictions as part of comprehensive tobacco control strategies. Previous research using data from Canadian provinces enacting such bans found more marked impacts on exposure in provinces with comprehensive strategies including discouraging smoking uptake and encouraging smoking cessation [8]. Recent evidence has also pointed to a role in exposure to smoking in cars in the incidence of asthma, which widens the potential health benefits of such legislation [10].

The ban is an example of the “Protect” element of the MPOWER policy approach to delivering the WHO Framework Convention on Tobacco Control programme to reduce the harms caused by smoking. Importantly, the purpose of the ban is to reduce child exposure to tobacco smoke, for which this study provides evidence, not to drive prosecutions (see, for example, coverage presenting the legislation as a failure due to the low number of arrests https://www.mirror.co.uk/news/uk-news/car-smoking-ban-massive-flop-10858407 (last visited 21^st^ August 2019)).

### Strengths and limitations

The strength of this natural experiment using high-quality data from Scotland and England is that because of its design the observed change can plausibly be ascribed to the policy intervention. A limitation is that there were only three data points for each country. Although survey data were not collected in precisely the same way between countries and years the approach we employed to harmonise measures was robust and any differences should not have affected changes over time. Exposure was based on self-report only, and reporting bias may have changed over time, although this would likely have been similar in both countries and so not significantly bias our findings. Interview dates were not available for the Scottish data and sampling in 2016 included almost two months after the introduction of their ban, resulting in some potential misclassification.

## Conclusions

Our results suggest that banning smoking in private vehicles carrying children has been successful in its main aim of reducing their exposure to tobacco smoke. Given children’s known vulnerability to second-hand smoke exposure, the observed reduction is likely to have resulted in improved health.

## Data Availability

Data for this study is available free of charge to UK university staff and students from the UK Data Archive

https://ukdataservice.ac.uk/

## APPENDIX

### Appendix contents

1. Further information on data sources
2. Appendix table 1 of study sample
3. Appendix table 2 of analyses in England only including children aged 11 – 15 years

### Further information on data sources

SDDU is a survey of children in school years 7 – 11 (aged 11-15 years) and is used to monitor the performance of the Government tobacco strategy [11][12][13][14][15].Data came from questionnaires administered to children at school in exam conditions and is designed to be representative of the gender, age, region and type of school in England. Data for Scotland come from the years 2012, 2014 and 2016, where children aged 13 years and older were asked to report their exposure to smoking in cars [16][17][18]. For children below this age, caregivers were asked to report exposure, but we have excluded this data due to concerns over the accuracy of caregiver reporting of exposure.

There were differences in the collection of data on markers of deprivation over time in England. In 2012 and 2014 children were asked if they received Free School Meals (FSM), but this measure was not used in 2016. The 2016 data used the Family Affluence Scale which asks children how many computers, vehicles and bathrooms their family has and assigned them a band from low to high [19]. We have harmonised these two measures by considered those receiving FSM or in the low FAS band as deprived. Scottish data used the Scottish Index of Multiple Deprivation as a marker of deprivation, and we harmonised the data by using the most deprived group as equivalent to receiving FSM or being in the lowest FAS band.

**Appendix table 1:** Study sample.

|  | England |  | Scotland |  |
| --- | --- | --- | --- | --- |
|  | Overall N | % exposed | Overall N | % exposed |
| 2012 | 4915 | 6.3 | 319 | 3.4 |
| 2014 | 3483 | 5.9 | 271 | 2.2 |
| 2016 | 6920 | 1.6 | 232 | 1.3 |
| Age 13 years | 5056 | 4.0 | 272 | 0.7 |
| Age 14 years | 4702 | 3.8 | 266 | 3.4 |
| Age 15 years | 5560 | 4.5 | 284 | 3.2 |
| Boys | 7617 | 3.3 | 429 | 1.4 |
| Girls | 7701 | 4.8 | 393 | 3.6 |
| Not deprived group | 13572 | 3.5 | 552 | 2.2 |
| Deprived group | 1746 | 8.8 | 270 | 3.0 |
| Total | 15318 | 4.1 | 822 | 2.4 |

**Appendix table 2:** Difference in difference analyses in England only including children aged 11 – 15 years.

|  | AOR | Lower CI | Upper CI |
| --- | --- | --- | --- |
| Regular exposure |  |  |  |
| England Trend | 1.03 | 0.88 | 1.21 |
| Policy Implementation | 0.26 | 0.18 | 0.36 |
| Monthly exposure |  |  |  |
| England Trend | 0.80 | 0.73 | 0.88 |
| Policy Implementation | 0.75 | 0.62 | 0.90 |
| Ever exposure |  |  |  |
| England Trend | 0.88 | 0.80 | 0.95 |
| Policy Implementation | 0.78 | 0.66 | 0.91 |

*These analyses use data from England only with a linear term for year (shown as “trend” 2012, 2014, 2016) and a binary variable to denote the one post-policy period (shown as “Policy Implementation”). Analyses are controlled for age (11 – 15 years, categorically), sex and a marker of deprivation and survey weights for non-response.*

